# Hybrid immunity improves the immune response after fourth Covid-19 vaccine dose in individuals with medical conditions predisposing to severe Covid-19

**DOI:** 10.1101/2023.12.21.23300374

**Authors:** Nina Ekström, Tuija M. Leino, Aapo Juutinen, Toni Lehtonen, Anu Haveri, Oona Liedes, Saimi Vara, Heini Salo, Arto A. Palmu, Hanna Nohynek, Timi Martelius, Merit Melin

## Abstract

**Background:** Data on Covid-19 booster vaccinations and subsequent infections on immune responses in the immunocompromised is limited. We studied antibody responses after the fourth dose and subsequent breakthrough infection to define patient groups benefiting most from boosters.

**Methods:** In Finland, fourth vaccine (booster) doses were first recommended for severely immunocompromised individuals, whom we invited to participate in 2022. We assessed spike protein specific IgG antibody levels and neutralizing antibodies (NAb) against the ancestral and Omicron BA.1 strains one month after the fourth dose from 488 adult participants and compared to the levels of 35 healthy controls after 3 doses. We used Bayesian generalized linear modelling to assess factors explaining antibody concentrations after the fourth dose. We assessed vaccine-induced and hybrid immunity six months after the last vaccine dose.

**Results:** Chronic kidney disease (CKD) and immunosuppressive therapy (IT) were identified as factors explaining sub-optimal antibody responses. The proportion of participants with a normal antibody response and NAbs were significantly lower in CKD patients as compared to controls. By the 6-month sampling one third of the participants became infected, which enhanced antibody levels notably in most immunocompromised participants.

**Conclusions:** Impaired antibody responses, especially NAbs against the Omicron lineage, predict limited protection in individuals with CKD, and highlight the need for alternative pharmaceutical preventive strategies. Vaccination strategies should take into account development of robust hybrid immunity responses also among the immunocompromised.

## Introduction

Immunocompromised individuals are at an increased risk for severe Coronavirus Disease 2019 (Covid-19) and at higher risk of mortality after Covid-19 hospitalization compared with immunocompetent individuals [1, 2]. The risk depends heavily on the extent to which the immune system has been suppressed [3].

Although vaccinations aim to protect these most vulnerable groups from severe Covid-19 disease, immunocompromised individuals have been shown to exhibit impaired responses to vaccines, including impaired or delayed antibody production [4, 5], diminished T cell responses in some studies [6, 7] and greater waning of immunity leading to waning vaccine effectiveness [8, 9] and they may not achieve the same level of protection as the immunocompetent [10, 11]. This may be due to immunomodulatory effects of the disease or condition itself or to treatment with immunosuppressive effects.

A primary series of two Covid-19 vaccine doses in immunocompromised populations generates impaired immune responses compared to the general population [12]. The World Health Organization therefore recommended a third dose for all immunocompromised persons [13]. In Finland, a fourth Covid-19 vaccine dose was recommended at a minimum interval of three months after the third dose in populations at risk for severe Covid-19 in late 2021 [14]. At the same time, a third (booster) dose was recommended for health care workers and older residents in long term care facilities, and subsequently for all adults [15]. Thereafter in 2022-23 subsequent booster doses have been recommended for those at greatest risk for severe Covid-19. However, specific information on which patient groups would benefit most from booster doses remains limited.

With the spread of Omicron variants, vaccinations have provided only very limited and short-lived protection from Covid-19 infections even in the boosted immunocompetent population. We have estimated, that 39% of the Finnish adult population had developed antibody-mediated hybrid immunity induced by a combination of vaccinations and infections during the first half of 2022, and over half by the end of 2022 [16]. Data on development of the hybrid immunity in different immunocompromised disease groups is scarce. With the continued evolution of new SARS-CoV-2 variants with increasing potential to evade immunity [17, 18], data on the persistence of vaccine induced and hybrid immunity is required to inform vaccination strategies in this high-risk population.

In this study we examined antibody levels after the fourth Covid-19 vaccine dose in a cohort of severely immunocompromised adults with varying underlying conditions. We aimed at identifying patient groups who could benefit from additional booster vaccinations the most. We used Bayesian modelling for evaluation of factors explaining diminished antibody responses. As the fourth vaccine dose in the immunocompromised individuals was considered their first booster dose, we compared their IgG and neutralizing antibody levels against the ancestral SARS-CoV-2 strain and Omicron BA.1 variant to levels in healthy controls after the third dose (booster). We additionally evaluated the persistence of vaccine and hybrid immunity induced antibodies up to six months after the fourth dose in immunocompromised individuals with different underlying conditions.

## Methods

### Study design and participants

The Covid-19 vaccine immunological study is a clinical trial conducted in Finland by the Finnish Institute for Health and Welfare (THL) within the wellbeing services counties of Uusimaa, Pirkanmaa, Kanta-Häme, Central Finland, Päijät-Häme and Ostrobothnia. No investigational products were used; the participants received Covid-19 vaccines in accordance with the national recommendations through routine healthcare service. For individuals with severe predisposing medical factors affecting the risk of Covid-19, a third vaccine dose had been recommended (on Sept 17, 2021) to supplement the primary vaccination series, at a minimum of two months after the second vaccine dose, but due to the short interval, this dose was not considered a booster [19]. For this group the recommendation of fourth Covid-19 vaccine dose at the minimum of three months following the previous dose was given on Dec 22, 2021 [20], and it was considered the first booster dose.

In Finland, a third vaccine dose was recommended (on Sept 21, 2021) at a minimum of six months after the primary vaccination series for healthcare workers and the elderly who had received the primary series doses with a short interval (<6 weeks)[21]. Later, on Nov 26, 2021, third vaccine doses were recommended for all healthcare workers at a minimum of six months following the second dose [22].

In January 2022, we invited 1440 subjects (aged ≥ 18 years) by random sampling to participate in the study. The inclusion criteria were previous receipt of three doses of a monovalent Covid-19 vaccine (BNT162b2, mRNA-1273 or ChAdOx1 nCoV-19) followed by a fourth monovalent vaccine dose (BNT162b2 or mRNA-1273) by Jan 25, 2022, and no previous registered SARS-CoV-2 infection. The subjects were invited by regular mail and were asked to donate a blood sample at their local healthcare district laboratory after the fourth dose. Any previous laboratory-confirmed SARS-CoV-2 infections were verified from register data. All study subjects were also asked on their first study visit whether they had previously received a positive result from an at-home antigen test. Seropositivity with a nucleoprotein IgG antibody test was additionally used to identify previous infections. Study subjects with a previous SARS-CoV-2 infection were excluded from this study (n=58).

To evaluate a normal vaccine response, we also recruited 65 healthcare workers at the Helsinki University Hospital (HUS) in December 2021 when they received their third Covid-19 vaccine. We selected to the control group all the healthcare workers (35/65) who had no predisposing medical factors affecting the risk of Covid-19 (as confirmed from healthcare registers), no known previous SARS-CoV-2 infection and who had received their first two vaccine doses (BNT162b2 or ChAdOx1 nCoV-19) at a dose interval >6 weeks (median 84, IQR 83-85 days). They received the third, i.e. booster dose (BNT162b2) 210 days (median, IQR 198-234) after the second dose, and provided post booster serum samples in January-February 2022.

We subsequently invited all study participants to a follow-up visit six months after the fourth or third dose, in May-July 2022 (Figure 1). The participants who had not received additional Covid-19 vaccinations between the first and second sampling were included in the analysis. The participants were considered to have hybrid immunity, if they had been infected with SARS-CoV-2 between the first and second study visit (at least seven days before second sampling).

**Figure 1.**
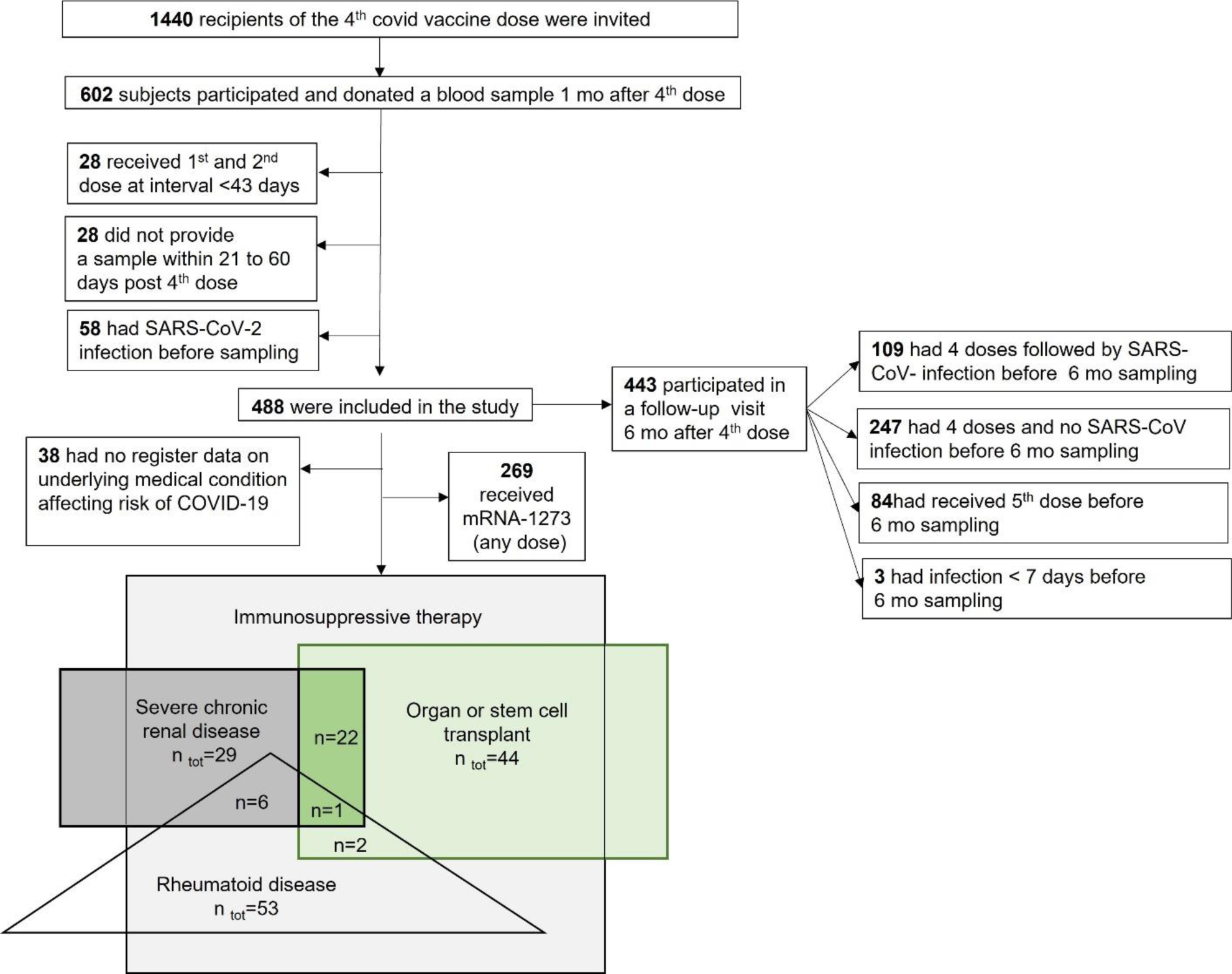
Study flow chart showing the immunocompromised participants included in the study. For analysis of immune responses by disease/treatment groups one month after the fourth vaccine dose we excluded participants who had received mRNA-1273 as the fourth dose. That was done because of better immunogenicity of mRNA-1273 suggested by the modelling data, the participants in the control groups did not receive mRNA-1273 (any dose) and limited register data on the dose of the mRNA-1273 vaccine (whole or half) used as fourth dose, which would not have allowed us to reliably compare different groups. For analysis of vaccine-induced and hybrid immunity (vaccine and infection) at six months after the fourth Covid-19 vaccine dose, we included participants who had not received additional Covid-19 vaccine doses and those who had a SARS-CoV-2 infection at least seven days before 6-month sampling. Because the proportions of those participants who had received mRNA-1273 vaccine (any doses) did not significantly differ in participants with vaccine or hybrid immunity at six months, we also included the participants who had received mRNA-1234 in the analysis of antibody levels at six months.

We retrieved information on Covid-19 vaccinations from the National Vaccination Register (THL), information on laboratory-confirmed SARS-CoV-2 infections from the National Infectious Diseases Register (THL), and information on predisposing factors for Covid-19 in the Care Register for Health Care (THL), the Register of Primary Health Care Visits (THL), the Special Reimbursement Register for Medicine Expenses and the Prescription Centre database (The Social Insurance Institution of Finland, KELA).

### Participant consent statement

The study was conducted according to the guidelines of the Declaration of Helsinki and approved by the National Committee on Medical Research Ethics (TUKIJA/347/2021) and the Finnish Medicines Agency Fimea as the regulatory authority (EudraCT 2021-004788-29). Written informed consent was obtained from all participants.

### Laboratory analyses

#### SARS-CoV-2 fluorescent multiplex immunoassay

We measured the concentration of serum IgG antibodies with an in-house fluorescent multiplex immunoassay (FMIA) [23] to SARS-CoV-2 nucleoprotein (N-IgG) and two spike protein (S-IgG) antigens; full-length spike protein (SFL) and receptor binding domain of spike protein (RBD). We used a threshold of 10 BAU/ml for N-IgG seropositivity resulting in 93% specificity and 100% sensitivity and thresholds 2 and 3 BAU/ml for SFL-IgG and RBD-IgG with 100% sensitivity and specificity, respectively. The assay has been calibrated to the WHO international standard [24].

#### Microneutralization assay

A live-virus cytopathic effect-based microneutralization assay (MNT) [25, 26] was performed to determine neutralizing antibody (NAb) titres against SARS-CoV-2 in subgroups of randomly selected samples (n=17-20/risk group). NAb titers were additionally determined for the first 27 samples taken of the 35 healthy controls. We used two SARS-CoV-2 viruses isolated in Finland between 2020-2022 representing the ancestral strain (WT) and the Omicron BA.1 subvariant [27]. Omicron BA.1 was the most prevalent variant in Finland from the end of 2021 to March 2022, followed gradually by Omicron BA.2 [28]. The isolation and propagation of the WT strain was performed in African green monkey kidney epithelial (Vero E6) cells [25], and of the BA.1 strain in VeroE6-TMPRSS2-H10 cells [29] and further propagated in Vero E6 cells for MNT. A tissue culture infectious dose 50% assay was performed for both viruses to achieve the comparable virus concentration among the different strains. Results were expressed as MNT titres; MNT titre ≥6 was considered positive, borderline positive when 4 and negative when <4. Borderline positive values were further confirmed by biological repeats.

#### Statistical methods

We assessed IgG concentrations in the control group one month after the third dose and used the 5^th^ percentile as the threshold for a normal response. To gain a more comprehensive insight into the variability and distribution of the normal response, we employed bootstrapping, generating 50,000 samples. This allowed us to estimate also the 95% highest density interval (HDI), providing a robust characterization of the response’s uncertainty and distribution.

Since most study participants had more than one immunosuppressive condition, a direct comparison of individual patient groups was not possible in this study setting. We therefore used modeling to help identify diseases and other factors that would explain low IgG concentrations following the fourth vaccine dose in the immunocompromised study population. We utilized a Bayesian gamma regression model with IgG concentrations as the response variable, and age, sex, the fourth vaccine product and the risk group status for various different predisposing conditions as explanatory variables. Assuming that only some of the explanatory variables truly impact the IgG concentrations, we used horseshoe priors [30] for the regression coefficients to induce sparsity in the model estimates. The model was applied separately for SFL- and RBD-IgG concentrations. The reference levels for categorical variables were defined as male for sex, BNT162b2 for the fourth vaccine product, and no risk factors for the risk group.

We calculated the geometric mean antibody concentrations (GMCs) and titers (GMTs) with 95% confidence intervals (CIs) for IgG and NAb levels, respectively, and compared the concentrations and titers of each group to control group, and the differences in antibody concentrations between vaccine and hybrid immunity groups with the Mann-Whitney test. MNT titers <4 were assigned a value of 2. We compared the proportions of subjects with normal antibody concentrations or MNT titer ≥4 with Fisher’s exact test.

Wilcoxon signed-rank test was used to compare antibody levels between different timepoints within groups. We calculated the Spearman correlation coefficient (ρ) and the statistical significance of the correlation between antibody concentrations and age, and between antibody concentrations and MNT titers. The statistical significance level of difference was set to P<0.05 and adjusted with Bonferroni correction to P<0.025-0.005, depending on the number of groups, time points and antibody/strain specificities in the comparison. Statistical analyses were performed with GraphPad v9 and R version 4.3.2.

## Results

### Characteristics of the participants who received 4 vaccine doses

In all, 602/1440 (42%) invited subjects participated (Figure 1). Of them 488 (81%) had received their first two vaccines (BNT162b2, ChAdOx1 nCoV-19 or mRNA-1273) with a dose interval of >6 weeks (IQR 83-84 days) followed by a third dose (BNT162b2, ChAdOx1 nCoV-19 or mRNA-1273) 112 days (median, IQR 90-128) after the second dose. They had received their fourth dose 106 days (median, IQR 99-112) after the third dose and provided their first sample for the study 36 days (median, IQR 32-41) after the fourth vaccine dose (BNT162b2 or mRNA-1273). We included these 488 participants to this study (Figure 1, Table 1). Half of the participants (264/488; 54%) received BNT162b2 and the other half (224/488, 46%) received mRNA-1273 as the fourth dose. The median age was 64 years (IQR 52-69) and 58.4% were female (Table 1). Most participants (443, 91%) attended a follow-up visit six months (median 5.6 months, IQR 5.4-5.9) after the fourth dose. Of the participants who had not received Covid-19 vaccines between the samplings (n=356), 109 (31%) experienced SARS-CoV-2 infection and provided post-infection samples for assessment of hybrid immunity (Table 1, Figure 4).

**Figure 4.**
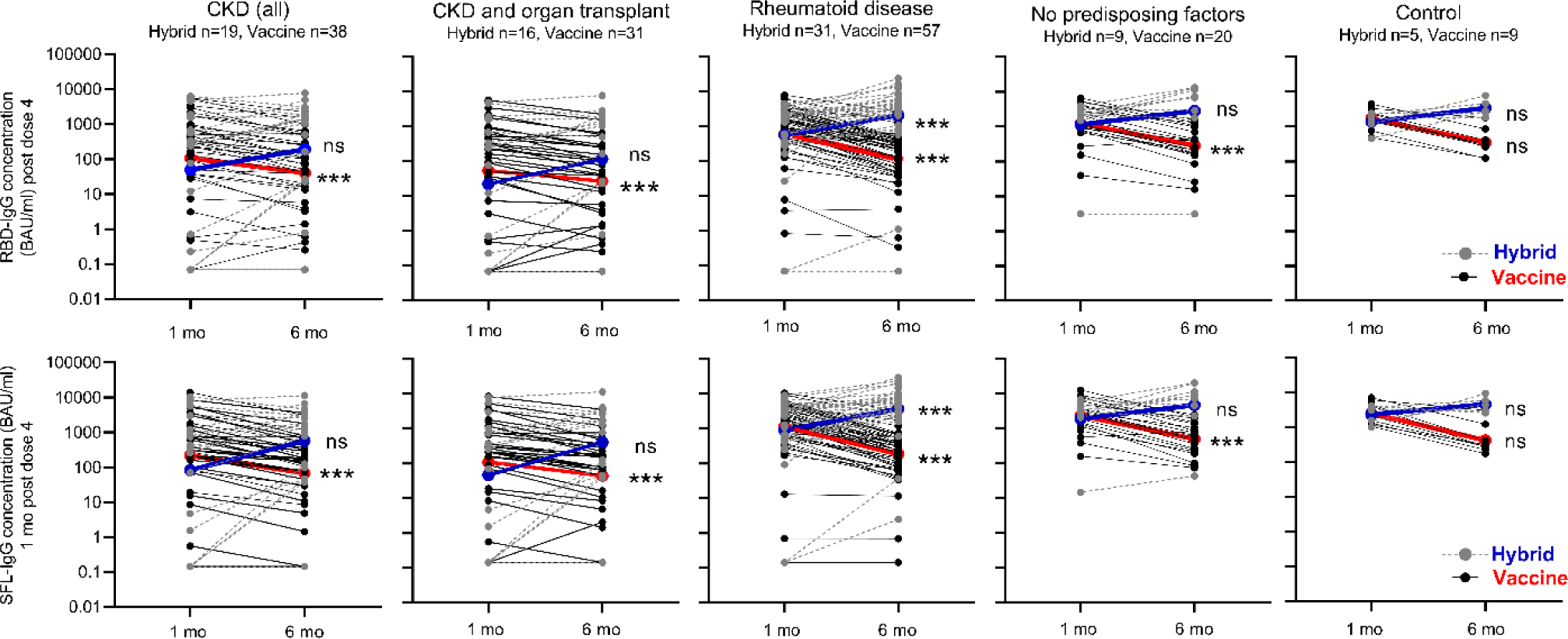
Kinetics of SFL- and RBD-IgG antibody concentrations (BAU/ml) in participants with hybrid or vaccine induced immunity by disease groups and in control group at one and six months after booster dose of Covid-19 vaccine. Participants who became infected during the 6-month follow-up and who thus developed hybrid immunity are shown by grey dotted lines. Participants with vaccine-induced immunity are shown by black solid lines. Geometric mean IgG concentrations of the groups are connected by blue (hybrid) and red (vaccine induced immunity) lines. Significant differences in mean antibody concentrations between one and six months following the last vaccine dose in participants with vaccine induced and hybrid immunity groups are shown; Wilcoxon signed-rank test, *P<0.005, **P<0.001, ***P<0.0001.

**Table 1.**
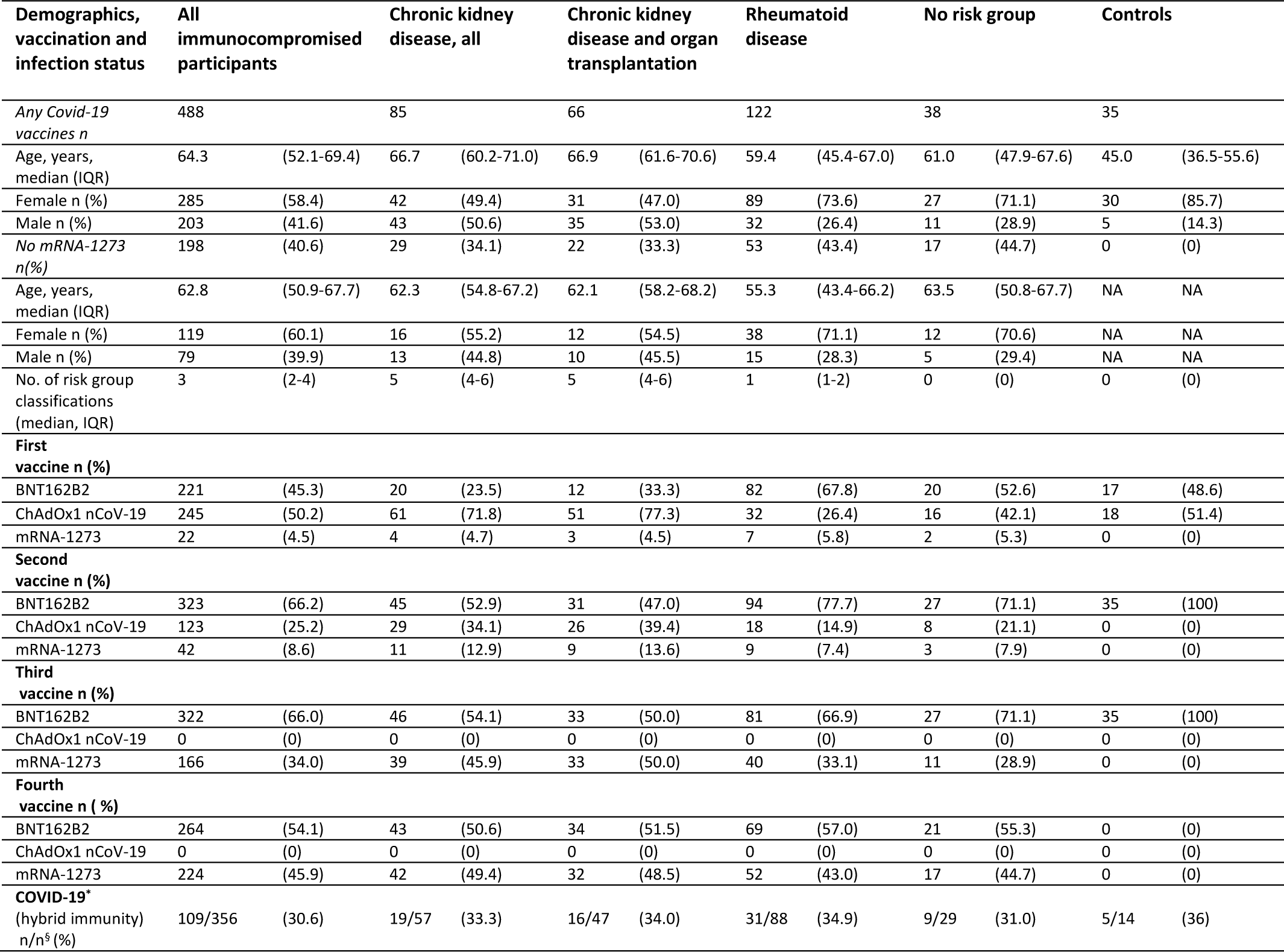
Baseline characteristics of the study participants.

Majority of the participants (394/488; 81%) had more than one (IQR 2-4) medical condition predisposing to severe Covid-19. The predisposing factors most frequently found were immunosuppressive therapy (IT)(70%), autoimmune diseases including rheumatoid diseases (42%), severe heart diseases (34%), cancer under treatment (31%), organ or stem cell transplant (23%), and severe chronic kidney disease (CKD)(17%) (Table 2). A small number (n=38) of the participants who had received four doses had no registered predisposing medical factors affecting the risk of Covid-19. These participants were included in the study as an additional control group.

**Table 2.** Posterior means and 95% credible i ntervals for regression coefficients from the Bayesian gamma regression models explaining the SFL- and RBD-IgG antibody concentrations 3 to 8 weeks after the fourth COVID-19 vaccine dose by age, gender, vaccine and predisposing factors. The reference levels for categorical variables were defined as male for sex, BNT162b2 for the product name of the 4th vaccination dose, and no risk factors for the risk group.

| Variable | N | Estimate<br>(SFL-IgG) | 95% CI<br>(SFL-IgG) | Estimate<br>(RBD-IgG) | 95% CI<br>(RBD-IgG) |
| --- | --- | --- | --- | --- | --- |
| Age (in years) |  | 0.995 | (0.985-1.003) | 0.995 | (0.984-1.003) |
| Sex -female |  | 1.001 | (0.874-1.169) | 1.001 | (0.876-1.159) |
| Vaccine – mRNA-1273 (fourth dose) | 224 | **1.568** | **(1.161-2.018)** | **1.510** | **(1.092-1.968)** |
| Cancer under active treatment | 149 | 1.088 | (0.954-1.534) | 1.098 | (0.963-1.606) |
| Autoimmune diseases* | 203 | 0.997 | (0.808-1.129) | 0.999 | (0.833-1.149) |
| Organ or stem cell transplant | 113 | 0.993 | (0.764-1.124) | 0.994 | (0.768-1.142) |
| Neurological disease or condition that interferes with breathing | 27 | 1.009 | (0.859-1.463) | 1.013 | (0.879-1.614) |
| Immunosuppressive therapy | 340 | **0.612** | **(0.455-0.845)** | **0.652** | **(0.482-0.940)** |
| Type 2 diabetes with drug therapy | 79 | 1.004 | (0.877-1.293) | 1.002 | (0.856-1.264) |
| Type 1 diabetes or adrenal insufficiency | 46 | 0.895 | (0.491-1.059) | 0.878 | (0.474-1.064) |
| Sleep apnea | 68 | 1.032 | (0.926-1.541) | 1.021 | (0.923-1.435) |
| Severe chronic pulmonary disease | 63 | 1.000 | (0.829-1.185) | 0.999 | (0.817-1.196) |
| Severe chronic liver disease | 19 | 1.001 | (0.798-1.393) | 1.006 | (0.838-1.498) |
| Severe chronic kidney disease | 85 | **0.479** | **(0.339-0.741)** | **0.490** | **(0.342-0.787)** |
| Severe disorders of the immune system | 21 | 0.991 | (0.631-1.177) | 0.994 | (0.633-1.222) |
| Severe heart diseases e.g heart failure | 166 | 0.990 | (0.771-1.127) | 0.989 | (0.754-1.108) |

\*Including diseases/conditions where immunosuppressive therapy is generally used (e.g rheumatoid diseases, psoriasis and inflammatory bowel disease)

### Characteristics of the healthy controls

The healthy controls who had received three vaccine doses provided a sample at 36 days (median, IQR 31-42) after the third dose (BNT162b2)(Table 1). The median age was 45 years (IQR 37-56) and 86% were female. Of them 14/35 (40%) participated in a 6-month follow-up visit, and five of them had infection between the samplings.

### Impact of different factors on antibody levels after the fourth dose

Since most participants who received the fourth vaccine dose had more than one medical condition predisposing to severe Covid-19 and the patient groups largely overlapped, we were not able to directly compare antibody responses between different patient groups in this study setting. We used Bayesian gamma regression model to assess which factors best explain low antibody concentrations after the fourth dose. The results presented in Table 2 show lowest estimates for chronic kidney disease (CKD) (0.479 [0.339-0.741]; 0.490[0.342-0.787]) and immunosuppressive therapy (IT) (0.612 [0.455-0.845]; 0.652 [0.482-0.940]) for SFL- and RBD-IgG antibodies, respectively.

Based on the modelling data, the effects of sex or age on antibody levels following the fourth dose were not significant. mRNA-1273 as the fourth dose (irrespectively of the previous vaccine products given) resulted in highest estimates (1.568 [1.161-2.018]; 1.510 [1.092-1.968]) suggesting better immunogenicity than of BNT162b2. However, since there was no information recorded in the National Vaccination Register on whether mRNA-1273 was given as a whole or half dose, we cannot reliably evaluate the immunogenicity of the mRNA-1273 compared to BNT162b2 when given as booster vaccine. The recommendation to halve the vaccine dose when mRNA-1273 was as used as the booster was given in October 2021 [31]. However, it is possible that, especially in those groups where the vaccine response was expected to be weaker, the full dose continued to be used. As a third dose, the full dose of either Covid-19 vaccine was recommended to the immunocompromised. However, since mRNA-1273 given as the fourth dose was associated with higher antibody levels compared to BNT162b2, and participants in the control group had not received mRNA-1273, we included only those immunocompromised participants who had received BNT162b2 as their fourth dose in the further analysis (Figure 1).

In this study, we focused further evaluation of IgG and neutralizing antibody levels on the group of CDK patients and those receiving IT. As with most participants, many of the participants living with CDK had comorbidities that affect immunocompetence. A total of 22/29 (76%) of participants living with CKD were organ transplant (OT) recipients. In this study we evaluated antibody levels in participants living with CKD or CKD with organ transplant (CDK with OT) but not CKD alone (only 7 subjects; Supplementary-Figure 1). We did not assess the group with OT, because of limited register data available on the time and type of the transplantation.

The IT group was large and heterogeneous, i.e. immunosuppressive treatment is combined with various diseases that also may weaken the immune response. Primarily, we would have selected participants receiving biological drugs (e.g anti-CD20 therapies such as Rituximab), as they are expected to affect antibody-mediated immune responses. However, as these drugs are administered in hospital, the information is not recorded in the registers to which we had access in this study (The Special Reimbursement Register for Medicine Expenses and the Prescription Centre database, The Social Insurance Institution of Finland, KELA). Since rheumatic patients formed one large group within the IT group, and biological drugs (including Rituximab) are used in the treatment of rheumatoid diseases (RD), we selected participants of this group to represent those receiving IT treatment (Figure 1).

### Antibody levels after the 3rd dose in healthy controls

The antibody concentrations in the healthy controls ranged between 338 and 4435 BAU/ml for RBD-IgG and 373 and 7036 BAU/ml for SFL-IgG, and the mean concentrations were 1414 BAU/ml (95% CI 1120-1784) and 2168 BAU/ml (95%CI 1713-2744), respectively (Figure 2). We determined the lower limit of a normal antibody response to vaccination as the 5^th^ percentile of the antibody concentrations; 398 and 716 BAU/ml for RBD-IgG and SFL-IgG, respectively. All controls had NAb against WT and Omicron BA.1; with GMTs 404 (95% CI 269-608) and 26 (95% CI 18-35), respectively (Figure 3).

**Figure 2.**
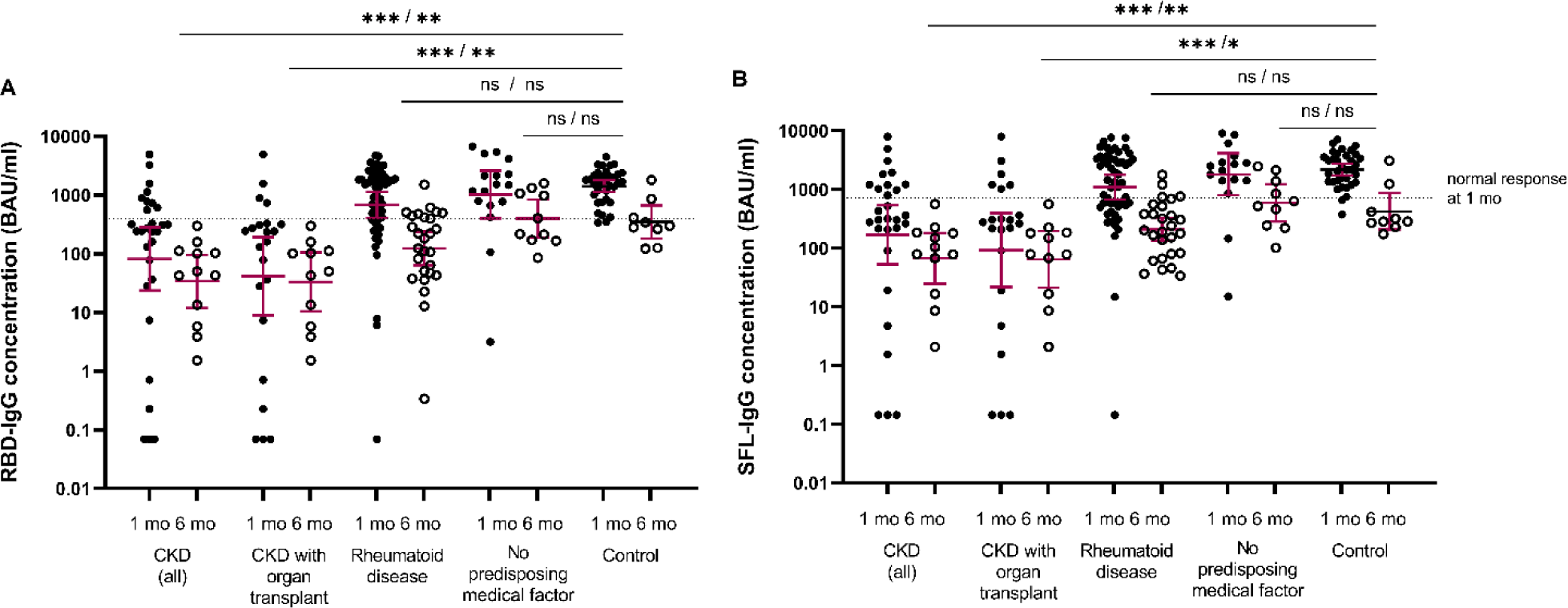
SARS-CoV-2 specific IgG antibody concentrations (GMC, 95% CI, BAU/ml) to receptor binding protein of Spike protein (RBD-IgG) (A) and full-length Spike protein (SFL-IgG) (B) in serum samples of participants with severe kidney disease (CKD, all) (n=29/13), CKD with organ transplant (N=22/11), rheumatoid disease (n=53/27), with no predisposing factors (n=17/10) and in healthy controls (n=35/9) at one/six months after Covid-19 vaccine, respectively. Participants who received mRNA-1273 vaccine are not included. 5th percentile of RBD- and SFL-IgG concentrations determined for the control group one months after third dose is shown as a threshold for a normal response (dashed line). Differences between each group and the control group were evaluated by Mann-Whitney test, *P<0.0025, **P<0.001, ***P<0.0001.

**Figure 3.**
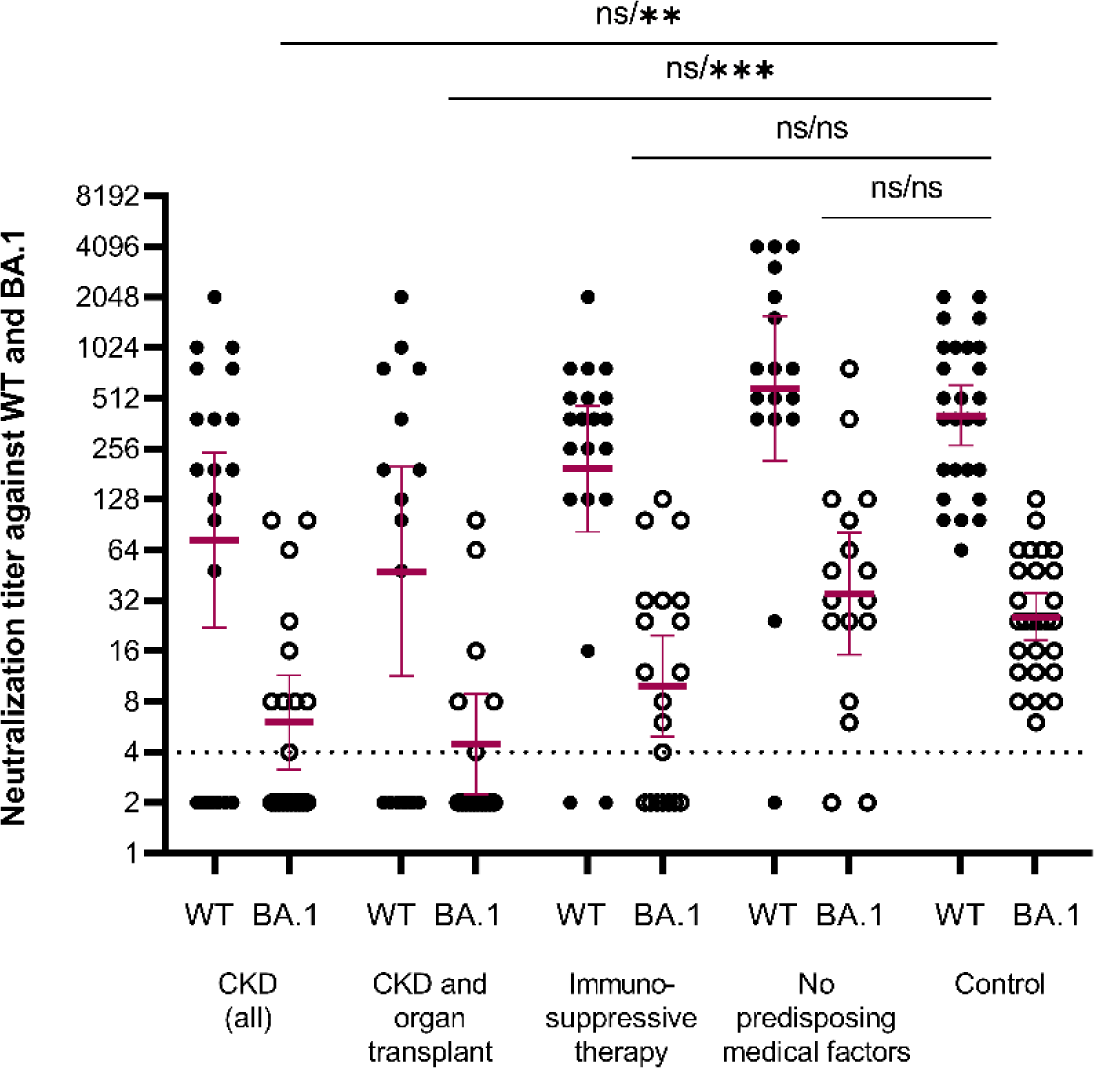
Neutralizing antibody titers against WT and BA.1 SARS-COV-2 strains one month after the fourth dose of Covid-19 vaccine in serum samples of participants with chronic kidney disease (CKD, all) (n=20), CKD and organ transplant (n=16), immunosuppressive therapy (n=20), with no predisposing factors (n=17) and in healthy controls (n=27). The threshold for a positive titer (>4) is shown as dashed line. Difference in NAb titers against WT and BA.1 strains between each group and the control group was evaluated by Mann-Whitney test, *P<0.005, **P<0.001, ***P<0.0001.

### Antibody levels and NAbs one month after the fourth dose by disease groups

Here we assessed the IgG antibody levels in the immunocompromised participants living with 1) CKD (with or without OT), 2) CKD with OT or 3) RD (with IT). We assessed the NAb levels in a small subgroup of participants living with 1) CKD (with or without OT), 2) CKD with OT and 3) those who received IT (without CKD, and with or without RD).

The majority (93%) of immunocompromised participants were seropositive for S-IgG antibodies after the fourth dose (Figure 2; Table 3). The percentage of participants with S-IgG antibody levels exceeding the lower limit of a normal response was significantly lower in the CKD and CKD with OT groups (31 and 18%) compared to healthy controls (P<0.001)(Table 3). In the CKD and CKD with OT groups the S-IgG antibody concentrations were significantly lower as compared to control group (P<0.0025; Figure 2). Within the CKD and CKD with OT groups, SFL- and RBD-concentrations significantly correlated negatively with age (ρ=- 0.405- -0.526, P<0.025), with the exception of RBD-IgG in the CKD with OT group.

**Table 3.**
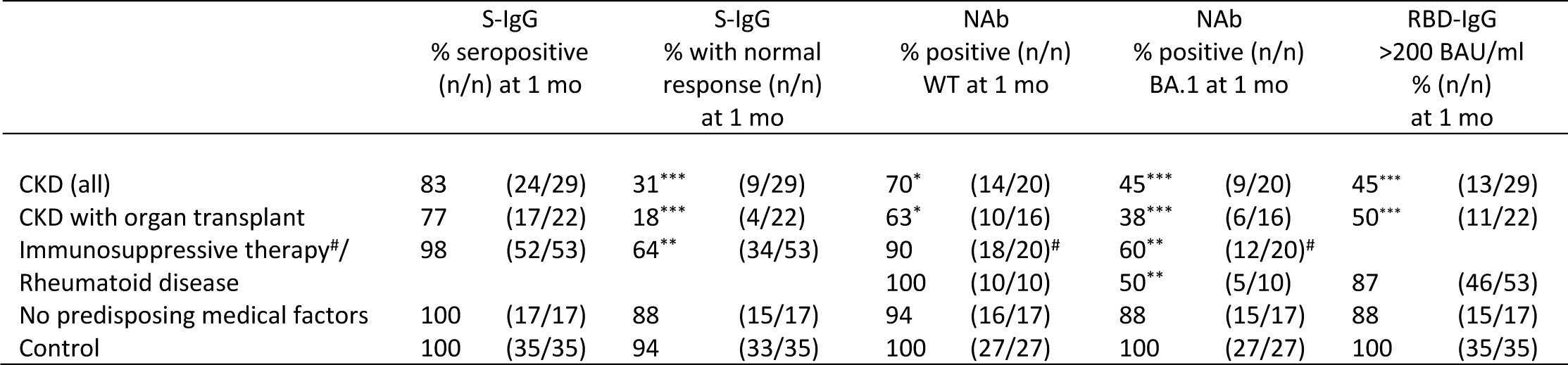
Percentages of seropositive participants, participants with a normal S-IgG response and NAbs, and participants with RBD-IgG specific IgG concentration >200 BAU/ml at one month after the booster dose by disease/control group of participants who had not received any dose of the mRNA-1273 vaccine. Of the participants who were on immunosuppressive therapy (IT) and had not received mRNA-1273 (n=112), 53/112 had rheumatoid disease (RD). We determined NAbs for 20/112 randomly selected participants with IT, and 10/20 of them also had RD. The tentative cut-off 200 BAU/ml was determined for CKD and IT groups as the minimum RBD-IgG concentration needed for positive neutralization titer against the Omicron BA.1 variant. Significant differences vs control group are shown; Fischer’s Exact test, *P<0.004, **P<0.001, ***P<0.0001.

We evaluated neutralizing antibodies for a randomly selected subgroup of participants with CKD (n=20, of whom with OT n=16) or IT (without CKD) (n=20, of whom with RD n=10). In the CKD, CKD with OT, and IT groups 70%, 63% and 90% of subjects had NAb against WT SARS-CoV-2 compared to 100% in the control group (Figure 3, Table 3). Mean NAb titers against BA.1 were >10-fold lower compared to WT in all groups. In the CKD, CKD with OT and IT groups 45%, 38% and 60% of participants had NAbs against BA.1 which was significantly less (P<0.004) than in the control group (100%)(Table 3). In the CKD and CKD with OT groups the mean NAb titers against BA.1 were significantly lower compared to the control group (P<0.001; Figure 3). Levels of NAbs against the WT and BA.1 strains correlated with the levels of SFL- and RBD-IgG antibodies (ρ=0.448-0.934; S-Figure 1). However, the minimum RBD-IgG antibody concentration generally required to neutralize the BA.1 variant was 200 BAU/ml in the CKD and IT groups. For samples with RBD-IgG concentrations below this, the NAb titer remained negative. In the control group all samples had RBD-IgG concentrations ≥ 338 BAU/ml and had NAbs against the BA.1 variant (Supplementary-Figure 1).

In the group of participants with four vaccine doses, but no registered medical factors affecting the risk of Covid-19, the mean NAb titers were comparable to titers in the control group after three doses of vaccine (Figure 3).

### Antibody levels six months after the booster dose in subjects with and without infection

During the follow-up, 109/356 (31%) of the immunocompromised participants experienced SARS-CoV-2 infection (Table 1). Of the infections, 68 (62%) were documented in the Infectious Diseases Register, and 42 (38%) were identified solely by >30% increase in N-IgG and/or S-IgG concentrations. Seven participants had hospital-treated COVID-19 disease, of which six had CKD (including four with CKD and OT), and one had cancer and severe heart disease. The median time from laboratory-confirmed SARS-CoV-2 infection to 6- month sampling was 76 days (range 12-181 days). Since the proportion of participants who received mRNA-1273 (any dose) was not significantly different in participants with or without an infection by six months after fourth dose (44 vs 71%; 44 vs 65%; 52 vs 52% and 56 vs 50%, for subjects with CKD, CKD with OT, RD and no risk group, respectively) we did not exclude mRNA-1273 recipients from the comparison of vaccine-induced and hybrid immunity. Only 14 of 35 participants in the control group participated in the 6-month sampling, and 5/14 were infected following the third vaccine dose (36%).

In participants without infection, the mean IgG concentrations decreased significantly in all groups; 2.0-3.2-fold in the CKD and CKD with OT, 5.8-6.1 -fold in the RD, and 4.9-6.0-fold in the control group (Figure 4).

The S-IgG concentrations were significantly lower in the CKD and CKD with OT than in the control group (P<0.0025, Figure 2).

After an infection, the mean S-IgG concentrations increased 4.7-9.0-fold in CKD and CKD with OT, 3.7-4.2 - fold in RD, and 1.9-2.5 -fold in the control group. The increase was significant (P<0.0001) in the RD group (Figure 4). Infection induced increases in RBD- and SFL-IgG concentrations in most subjects with CKD (82 and 91%), CKD with OT (69 and 81%) or RD (69 and 75%), respectively (Figure 4). In the control group, the RBD- and SFL-IgG concentrations increased after infection in 3 of the 5 participants. Six months after the fourth vaccine dose, RBD- an SFL-IgG concentrations were significantly higher in participants who had an infection, (i.e. those who had developed hybrid immunity during the follow-up) compared to those without infection(P<0.005), except for CKD with OT group, and RBD-IgG in the CKD group (Figure 4).

## Discussion

In this study we show that the magnitude of the antibody responses after the fourth vaccine dose were sub-optimal particularly in participants with severe CKD. On the other hand, we showed that infections enhanced antibody levels in most immunocompromised participants. With the emergence of the Omicron variant and the variants that followed, infections have been widespread also in the vaccinated population.

Hybrid immunity has been found to provide higher and more durable protection against reinfection and severe disease than vaccination alone [32]. The data on immune response to the fourth dose/booster doses and hybrid immunity is limited, especially in disease-specific subgroups e.g individuals living with CKD or RD. Our study showed that after the fourth dose the proportion of participants with a normal antibody response and those with NAb against the Omicron BA.1 variant were significantly lower in individuals with CKD or RD as compared to healthy controls. Impaired neutralizing antibody responses predict limited protection. We additionally showed that despite impaired responses to booster vaccination, infections boosted the antibody concentrations from what had been achieved with the fourth vaccine dose in the majority of the immunocompromised participants. Further, we conclude that mRNA-1273, as the fourth dose, was associated with higher immunogenicity as compared to BNT162b2 in immunocompromised subjects.

CKD and RD associated with immunosuppressive therapy have also in previous studies shown to impact immune responses to Covid-19 vaccination. Previous studies have shown overall seroconversion rates as high as in healthy controls after primary vaccination [33–35], but significantly lower mean RBD-IgG antibody levels, NAb antibody titers (against wt) and lower T-cell responses compared to healthy subjects [6, 33, 36–38]. The third vaccine dose given more than three months after the primary vaccination has been shown in patients with RD to increase the RBD-IgG level, NAb titer against wt/Omicron and T cell response significantly, and compared to healthy controls lower or similar response levels have been reported [37, 39]. In patients with CKD, the third dose has been demonstrated to increase the antibody levels, proportion of participants with NAbs and NAb titer levels and T cell responses [40–43] and seroconversion has been observed in patients who did not respond after two vaccine doses [34, 44]. However, in kidney transplant recipients the rates of response have remained suboptimal [41]. Due to significant waning of antibody levels in the months following the third dose [38, 44, 45] and the emergence of SARS-CoV-2 variants with a high potential of immune escape The European Center for Disease Prevention and Control and European Medicine Agency recommended in July 2022 the administration of the fourth dose to people above 60 years as well as vulnerable persons of any age [46]. Fourth vaccine dose has been shown to elicit higher antibody levels than after the third dose in immunocompromised patients [47], but lower levels than in controls after three doses in studies with patients on immunosuppressive therapy [48], RD [49] and with CKD [40, 44]. Few studies have, however, reported NAbs, T cell mediated or hybrid immunity and persistence of antibodies after fourth dose in patients with CKD and RD. One study reported increased neutralizing activity and T cell response after fourth dose in patients with RD, with only a minority displaying a level of neutralization above baseline for the Omicron BA.2 strain [49].

We report here significantly impaired antibody levels and NAb titers against BA.1 strain after the fourth dose in participants with CKD and often with OT. This finding suggests that even after a booster dose the antibody-based immunity is limited in individuals living with CKD, and even more limited in those living with CKD and OT. Progressive waning of antibody levels during the months after the fourth dose shown in this study and emergence of SARS-CoV-2 variants with a high potential of immune escape highlight the need for booster vaccinations for people living with CKD. Despite lower humoral responses vaccination can significantly reduce the odds of severe disease, hospitalization and death in patients with CKD. However, for CKD patients with persistent poor vaccine responses, and especially for those with CKD and OT, alternative preventive strategies such as new adapted vaccines or a combined approach of immunization, pre-exposure prophylaxis and early post-exposure treatment using direct-acting antivirals and neutralizing monoclonal antibodies may be an option.

Previous data on antibody responses to breakthrough infection in specific immunocompromised patient groups, especially after fourth vaccine dose is scarce. Previous studies have reported higher antibody levels after hybrid compared to vaccine induced immunity after two or three vaccine doses in patients with CKD [44, 50] and immune-mediated inflammatory diseases [48]. We show in this study that despite impaired antibody responses to the fourth dose, most immunocompromised participants were able to mount a substantial antibody response to infection. At the time of the follow-up of our study (Feb-June 2022) molecular testing was not routinely performed in relation to infections with mild symptoms. Instead, at-home antigen tests were recommended. This practice is reflected in the relatively large proportion of participants (32%) whose breakthrough infection was detected solely by antibody measurement. Due to high overall attendance at the follow-up visit (91%) it is unlikely, that participants with a known or suspected infection were over-represented. During the follow-up 7/109 (6%) of infections in all participants and 6/19 (32%) in participants with CKD resulted in hospital admission. Our data demonstrates that also a relatively mild or asymptomatic infection is able to function as a booster in vaccinated immunocompromised individuals.

The strengths of our study include evaluation of factors affecting immune responses to booster vaccination in immunocompromised participants with variable underlying conditions. Also, we compared immune responses to a group of healthy controls with matched vaccine products and similar schedule for sampling. Because of the missing register data on the used dose of the fourth mRNA-1273 vaccine (whole or half) and the greater immunogenicity of mRNA-1273 in comparison to BNT162b2 shown in this study, we performed the comparisons between disease groups after excluding participants who had received mRNA-1273. In addition, we assessed immune responses by antibody levels and NAbs against WT and the prevalent variant at the time of sampling, Omicron BA.1. Neutralizing antibodies are considered as a more relevant correlate of protection than antibody levels against severe disease [51, 52]. In addition to detecting previous SARS-CoV-2 infections by molecular tests we also performed N-IgG antibody measurements and asked the participants about coronavirus at-home antigen tests to minimize the possibility of previous infections influencing our results.

Our study evaluated humoral immune response to vaccination and infection, but we did not assess cellular immunity, which is a limitation in our study. Recent data shows that despite impaired B-cell responses to vaccination specific T-cell responses are less dependent on disease or treatment characteristics [37, 39, 49, 53]. The ability to mount a T-cell response depends essentially on the extent to which immunodeficiency or immunosuppressive therapy affect T cells. Subjects with primary immune deficiencies were found to have robust T cell responses which can mediate protection against severe COVID-19 disease [53, 54]. The register-based data used in our study was partly limited as not all data on immunosuppressive treatments given as specialized medical treatment were available e.g B-cell depleting therapies such as anti-CD20 or immunosuppressive drug mycophenolate mofetil which have been shown to negatively affect antibody responses [37]. We defined the study subgroups solely by the underlying disease (ICD-10 codes) notwithstanding the data from previous studies suggesting that the immune status rather than the disease itself is responsible for impaired immune responses [37, 55]. Since most participants had more than one predisposing medical factor and the groups largely overlapped a direct comparison between patient groups was not possible, which is also a limitation. During the ongoing follow-up of the study, we will assess T cell responses and extend the register-based data.

The exposure of different population groups to infections may have varied, especially during the first epidemic waves, when containment measures were widely used. The proportion of immunocompromised participants who became infected (31%) during the first Omicron wave during this study, was, however, similar to the age-standardized seroprevalence of the population (39 %) in the Finnish population study carried out at the same time [16]. Considering that we excluded those participants who were infected before receiving the fourth vaccine dose, the cumulative seroprevalence in the immunocompromised participants would probably have been even closer to the general population.

Fourth dose of an mRNA vaccine given to immunocompetent individuals four months after the third dose was shown to restore the antibody concentrations to the level seen after the third dose, but not significantly boost the concentrations [56]. A longer interval between vaccine doses, or between vaccination and infection, is recognised to increase immunogenicity [57, 58]. Very long intervals, up to at least 400 days, between vaccination and infection were shown to result in not only stronger antibody responses but also better cross-neutralization of different Omicron variants. In our study, the subjects had infection after a short time, within six months of the fourth vaccine dose. Despite this, the infection notably enhanced the antibody responses beyond the levels measured after the fourth dose. Further booster doses have been recommended for immunocompromised subjects, and some may have received up to eight doses by the end of 2023.

Long-term benefits of continued boosting of immunocompromised subjects remain to be seen. In the current epidemiological situation, after many epidemic waves, most people have been infected, and many even several times. In the light of the current information, it should be critically evaluated whether booster doses are useful to be given repeatedly at short intervals to those immunocompromised or whether vaccinations should be scheduled closer to the start of the expected epidemic wave e.g in the autumn season.

## Acknowledgements

We thank all the participants of the study, and Marja Leinonen, Arja Rytkönen, Mervi Lasander, Janne Salimäki and Pirjo Tarkiainen for sample and consent handling, Leena Saarinen, Marja Suorsa, Lotta Hagberg, Saara Suopanki, Raisa Hanninen for laboratory analyses, Anna Solastie and Camilla Virta for laboratory analyses and assay development, Joni Niemi and Anita Nieminen for sample management, Esa Ruokokoski, Juha Oksanen, Dennis Ahlfors, Mikko Aura, Tommi Korhonen for data management, study coordinators and investigators Elina Isosaari, Ritva Syrjänen, Heta Nieminen, Päivi Siren and Maila Kyrölä, the Study Steering committee Terhi Kilpi, Jukka Jokinen, Carita Savolainen-Kopra, Simopekka Vänskä, Jaana Halonen, Otto Helve and Mia Kontio for participation in the clinical study design, Erika Lindh and Niko Tervo for providing SARS-CoV-2 variant surveillance data, the countless others who participated in sequencing and sample collection all over Finland.

## Author contributions

Conceptualization, M.M., A.A.P. T.M.L and H.N; Methodology, T.L., A.J, N.E; Software,T.L; A.J; Validation, T.L., A.J, N.E., A.H.,O.L.; Formal Analysis, T.L, A.J, N.E.,O.L; Investigation, T.L., A.J.,N.E., A.H., O.L., S.V; Resources, M.M., A.A.P. and H.N; Data Curation, T.L., A.J, N.E, O.L; Writing – Original Draft Preparation, N.E.,M.M., A.J.,T.O.L; Writing – Review & Editing, all authors; Visualization, N.E; Supervision, M.M., A.A.P. and H.N; Project Administration, M.M., N.E. and H.N.; Funding Acquisition, M.M., A.A.P. and H.N .

## Data Availability Statement

The data presented in this study is available on a reasonable request from the corresponding author.

## References

1. Williamson EJ, Walker AJ, Bhaskaran K, et al. Factors associated with COVID-19-related death using OpenSAFELY. Nature 2020;584:430–6.

2. Semenzato L, Botton J, Drouin J, et al. Characteristics associated with the residual risk of severe COVID-19 after a complete vaccination schedule: A cohort study of 28 million people in France. Lancet Reg Health Eur 2022;19:100441.

3. Evans RA, Dube S, Lu Y, et al. Impact of COVID-19 on immunocompromised populations during the Omicron era: insights from the observational population-based INFORM study. Lancet Reg Health Eur 2023;35:100747.

4. Lee, Ainsley Ryan Yan Bin, Wong SY, Chai LYA, et al. Efficacy of covid-19 vaccines in immunocompromised patients: systematic review and meta-analysis. BMJ 2022;376:e068632–068632.

5. Widhani A, Hasibuan AS, Rismawati R, et al. Efficacy, Immunogenicity, and Safety of COVID-19 Vaccines in Patients with Autoimmune Diseases: A Systematic Review and Meta-Analysis. Vaccines (Basel) 2023;11:1456. doi: 10.3390/vaccines11091456.

6. Ehmsen S, Asmussen A, Jeppesen SS, et al. Antibody and T cell immune responses following mRNA COVID-19 vaccination in patients with cancer. Cancer Cell 2021;39:1034–6.

7. Sanders JF, Messchendorp AL, de Vries RD, et al. Antibody and T-Cell Responses 6 Months After Coronavirus Disease 2019 Messenger RNA-1273 Vaccination in Patients With Chronic Kidney Disease, on Dialysis, or Living With a Kidney Transplant. Clin Infect Dis 2023;76:e188–99.

8. Andrews N, Stowe J, Kirsebom F, et al. Effectiveness of COVID-19 booster vaccines against COVID-19-related symptoms, hospitalization and death in England. Nat Med 2022;28:831–7.

9. Andrews N, Tessier E, Stowe J, et al. Duration of Protection against Mild and Severe Disease by Covid-19 Vaccines. N Engl J Med 2022;386:340–50.

10. Marra AR, Kobayashi T, Suzuki H, et al. The effectiveness of coronavirus disease 2019 (COVID-19) vaccine in the prevention of post-COVID-19 conditions: A systematic literature review and meta-analysis. Antimicrob Steward Healthc Epidemiol 2022;2:e192.

11. Tan TT, Ng HJ, Young B, et al. Effectiveness of vaccination against SARS-CoV-2 and the need for alternative preventative approaches in immunocompromised individuals: a narrative review of systematic reviews. Expert Rev Vaccines 2023;22:341–65.

12. Boyarsky BJ, Werbel WA, Avery RK, et al. Antibody Response to 2-Dose SARS-CoV-2 mRNA Vaccine Series in Solid Organ Transplant Recipients. JAMA 2021;325:2204–6.

13. Interim recommendations for an extended primary series with an additional vaccine dose for COVID-19 vaccination in immunocompromised persons. Available at: https://www.who.int/publications/i/item/WHO-2019-nCoV-vaccines-SAGE_recommendation-immunocompromised-persons. Accessed Dec 21, 2023.

14. Salo H, Lehtonen T, Auranen K, Baum U, Leino T. Predictors of hospitalisation and death due to SARS-CoV-2 infection in Finland: A population-based register study with implications to vaccinations. Vaccine 2022;40:3345–55.

15. THL recommends that third doses of the corona vaccine start to be offered to all 18-year-olds 5-6 months after the second dose. Available at: https://thl.fi/-/thl-suosittelee-etta-kolmansia-koronarokoteannoksia-aletaan-tarjota-kaikille-18-vuotta-tayttaneille-5-6-kuukautta-toisesta-annoksesta. Accessed Dec 20, 2023.

16. Solastie A, Nieminen T, Ekström N, et al. Changes in SARS-CoV-2 seroprevalence and population immunity in Finland, 2020-2022. Emerg Microbes Infect 2023;12:2222849.

17. Kurhade C, Zou J, Xia H, et al. Low neutralization of SARS-CoV-2 Omicron BA.2.75.2, BQ.1.1 and XBB.1 by parental mRNA vaccine or a BA.5 bivalent booster. Nat Med 2023;29:344–7.

18. Wang Q, Iketani S, Li Z, et al. Alarming antibody evasion properties of rising SARS-CoV-2 BQ and XBB subvariants. Cell 2023;186:279,286.e8.

19. THL instructs municipalities to offer third vaccine doses to people with severe immune deficiency. Available at: https://thl.fi/-/thl-ohjeistaa-kuntia-tarjoamaan-kolmansia-rokoteannoksia-voimakkaasti-immuunipuutteisille. Accessed Dec 21, 2023.

20. THL recommends fourth vaccine doses to people with severe immune deficiency . Available at: https://thl.fi/fi/-/thl-suosittelee-vakavasti-immuunipuutteisille-neljatta-koronarokoteannosta. Accessed Dec 21, 2023.

21. THL has instructed municipalities on the 3rd corona vaccine doses for those vaccinated with a short dose interval - the third dose not yet for men under 30. Available at: https://thl.fi/-/thl-ohjeistanut-kuntia-lyhyella-annosvalilla-rokotettujen-3.-koronarokoteannoksista-kolmatta-annosta-ei-viela-alle-30-vuotiaille-miehille. Accessed Dec 21, 2023.

22. THL recommends third doses of covid-19vaccine for those treating corona patients and other critical care staff. Available at: https://thl.fi/-/thl-suosittelee-kolmansia-koronarokoteannoksia-koronapotilaita-hoitaville-seka-muulle-kriittisen-hoidon-henkilokunnalle. Accessed Dec 20, 2023.

23. Solastie A, Virta C, Haveri A, et al. A Highly Sensitive and Specific SARS-CoV-2 Spike- and Nucleoprotein-Based Fluorescent Multiplex Immunoassay (FMIA) to Measure IgG, IgA, and IgM Class Antibodies. Microbiol Spectr 2021;9:e0113121,21. Epub 2021 Nov 17.

24. Establishment of the WHO international standard and reference panel for anti-SARS-CoV-2 antibody. WHO/BS/20202402. Available at: https://www.who.int/publications/m/item/WHO-BS-2020.2403. Accessed Dec 21, 2023.

25. Haveri A, Smura T, Kuivanen S, et al. Serological and molecular findings during SARS-CoV-2 infection: the first case study in Finland, January to February 2020. Euro Surveill 2020;25:2000266. doi: 10.2807/1560.

26. Haveri A, Ekström N, Solastie A, et al. Persistence of neutralizing antibodies a year after SARS-CoV-2 infection in humans. Eur J Immunol 2021;51:3202–13.

27. Haveri A, Solastie A, Ekström N, et al. Neutralizing antibodies to SARS-CoV-2 Omicron variant after third mRNA vaccination in health care workers and elderly subjects. Eur J Immunol 2022;52:816–24.

28. Genomic surveillance of SARS-CoV-2. Available at: https://thl.fi/en/web/infectious-diseases-and-vaccinations/what-s-new/coronavirus-covid-19-latest-updates/coronavirus-variants/genomic-surveillance-of-sars-cov-2. Accessed Dec 21, 2023.

29. Rusanen J, Kareinen L, Szirovicza L, et al. A Generic, Scalable, and Rapid Time-Resolved Förster Resonance Energy Transfer-Based Assay for Antigen Detection-SARS-CoV-2 as a Proof of Concept. mBio 2021;12:e00902–21.

30. Handling Sparsity via the Horseshoe. 12th International Conference on Articial Intelligence and Statistics (AISTATS) 2009, Clearwater Beach, Florida, USA. 2009 Clearwater Beach, Florida, USA: p. Volume 5 of JMLR: W&CP 5.

31. THL recommends offering the third dose of the corona vaccine to people over 60 and medical risk groups. Available at: https://thl.fi/-/thl-suosittelee-kolmannen-koronarokoteannoksen-tarjoamista-60-vuotta-tayttaneille-ja-laaketieteellisille-riskiryhmille. Accessed Dec 21, 2023.

32. Bobrovitz N, Ware H, Ma X, et al. Protective effectiveness of previous SARS-CoV-2 infection and hybrid immunity against the omicron variant and severe disease: a systematic review and meta-regression. Lancet Infect Dis 2023;23:556–67.

33. Sanders JF, Bemelman FJ, Messchendorp AL, et al. The RECOVAC Immune-response Study: The Immunogenicity, Tolerability, and Safety of COVID-19 Vaccination in Patients With Chronic Kidney Disease, on Dialysis, or Living With a Kidney Transplant. Transplantation 2022;106:821–34.

34. Bouwmans P, Messchendorp AL, Imhof C, et al. Impact of immunosuppressive treatment and type of SARS-CoV-2 vaccine on antibody levels after three vaccinations in patients with chronic kidney disease or kidney replacement therapy. Clin Kidney J 2022;16:528–40.

35. Simader E, Tobudic S, Mandl P, et al. Importance of the second SARS-CoV-2 vaccination dose for achieving serological response in patients with rheumatoid arthritis and seronegative spondyloarthritis. Ann Rheum Dis 2022;81:416–21.

36. Mandl P, Tobudic S, Haslacher H, et al. Response to SARS-CoV-2 vaccination in systemic autoimmune rheumatic disease depends on immunosuppressive regimen: a matched, prospective cohort study. Ann Rheum Dis 2022;81:1017–22.

37. Corradini P, Agrati C, Apolone G, et al. Humoral and T-Cell Immune Response After 3 Doses of Messenger RNA Severe Acute Respiratory Syndrome Coronavirus 2 Vaccines in Fragile Patients: The Italian VAX4FRAIL Study. Clin Infect Dis 2023;76:e426–38.

38. Tobudic S, Simader E, Deimel T, et al. The accelerated waning of immunity and reduced effect of booster in patients treated with bDMARD and tsDMARD after SARS-CoV-2 mRNA vaccination. Front Med (Lausanne) 2023;10:1049157.

39. Kartnig F, Mrak D, Simader E, et al. Safety and immunogenicity of a third COVID-19 vaccination in patients with immune-mediated inflammatory diseases compared with healthy controls. Ann Rheum Dis 2023;82:292–300.

40. Tillmann F, Figiel L, Ricken J, et al. Effect of Third and Fourth mRNA-Based Booster Vaccinations on SARS-CoV-2 Neutralizing Antibody Titer Formation, Risk Factors for Non-Response, and Outcome after SARS-CoV-2 Omicron Breakthrough Infections in Patients on Chronic Hemodialysis: A Prospective Multicenter Cohort Study. J Clin Med 2022;11:3187. doi: 10.3390/jcm11113187.

41. Poli MC, Vial C, Rey-Jurado E, et al. A Third Dose of SARS-CoV-2 mRNA Vaccine Improves Immune Response in Chronic Kidney Disease Patients. Vaccines (Basel) 2023;11:1012. doi: 10.3390/vaccines11051012.

42. Westhoff TH, Seibert FS, Anft M, et al. A third vaccine dose substantially improves humoral and cellular SARS-CoV-2 immunity in renal transplant recipients with primary humoral nonresponse. Kidney Int 2021;100:1135–6.

43. Quiroga B, Soler MJ, Ortiz A, et al. Humoral Response to Third Dose of SARS-CoV-2 Vaccines in the CKD Spectrum. Clin J Am Soc Nephrol 2022;17:872–6.

44. Quiroga B, Soler MJ, Ortiz A, et al. Humoral response after the fourth dose of the SARS-CoV-2 vaccine in the CKD spectrum: a prespecified analysis of the SENCOVAC study. Nephrol Dial Transplant 2023;38:969–81.

45. Bertrand D, Lemée V, Laurent C, et al. Waning antibody response and cellular immunity 6 months after third dose SARS-Cov-2 mRNA BNT162b2 vaccine in kidney transplant recipients. Am J Transplant 2022;22:1498–500.

46. ECDC and EMA update recommendations on additional booster doses of COVID-19 vaccines. Available at: https://www.ecdc.europa.eu/en/news-events/ecdc-and-ema-update-recommendations-additional-booster-doses-covid-19-vaccines. Accessed Dec 21, 2023.

47. Martinelli S, Pascucci D, Laurenti P. Humoral response after a fourth dose of SARS-CoV-2 vaccine in immunocompromised patients. Results of a systematic review. Front Public Health 2023;11:1108546.

48. Bjørlykke KH, Ørbo HS, Tveter AT, et al. Four SARS-CoV-2 vaccine doses or hybrid immunity in patients on immunosuppressive therapies: a Norwegian cohort study. Lancet Rheumatol 2023;5:e36–46.

49. Rescigno M, Agrati C, Salvarani C, et al. Neutralizing antibodies to Omicron after the fourth SARS-CoV-2 mRNA vaccine dose in immunocompromised patients highlight the need of additional boosters. Front Immunol 2023;14:1104124.

50. Nazaruk P, Tkaczyk I, Monticolo M, et al. Hybrid Immunity Provides the Best COVID-19 Humoral Response in Immunocompromised Patients with or without SARS-CoV-2 Infection History. Vaccines (Basel) 2023;11:1380. doi: 10.3390/vaccines11081380.

51. Gilbert PB, Montefiori DC, McDermott AB, et al. Immune correlates analysis of the mRNA-1273 COVID-19 vaccine efficacy clinical trial. Science 2022;375:43–50.

52. Khoury DS, Cromer D, Reynaldi A, et al. Neutralizing antibody levels are highly predictive of immune protection from symptomatic SARS-CoV-2 infection. Nat Med 2021;27:1205–11.

53. Hurme A, Jalkanen P, Marttila-Vaara M, et al. T cell immunity following COVID-19 vaccination in adult patients with primary antibody deficiency - a 22-month follow-up. Front Immunol 2023;14:1146500.

54. van Leeuwen, Leanne P M, Grobben M, GeurtsvanKessel CH, et al. Immune Responses 6 Months After mRNA-1273 COVID-19 Vaccination and the Effect of a Third Vaccination in Patients with Inborn Errors of Immunity. J Clin Immunol 2023;43:1104–17.

55. Azzolini E, Pozzi C, Germagnoli L, et al. mRNA COVID-19 vaccine booster fosters B- and T-cell responses in immunocompromised patients. Life Sci Alliance 2022;5:e202201381. doi: 10.26508/lsa.202201381. Print 2022 Jun.

56. Regev-Yochay G, Gonen T, Gilboa M, et al. Efficacy of a Fourth Dose of Covid-19 mRNA Vaccine against Omicron. N Engl J Med 2022;386:1377–80.

57. Parry H, Bruton R, Stephens C, et al. Extended interval BNT162b2 vaccination enhances peak antibody generation. NPJ Vaccines 2022;7:14-w.

58. Bates TA, Leier HC, McBride SK, et al. An extended interval between vaccination and infection enhances hybrid immunity against SARS-CoV-2 variants. JCI Insight 2023;8:e165265. doi: 10.1172/jci.insight.165265.

