## Supplementary file for "Hybrid immunity improves the immune response after fourth Covid-19 vaccine dose in individuals with medical conditions predisposing to severe Covid-19"

### Supplementary material

**Supplementary-Figure 1.** The correlation between RBD-IgG and SFL-IgG antibody concentrations (BAU/ml) and neutralizing antibody titers against wild type (WT) and Omicron BA.1 SARS-CoV-2 strains in samples taken one month after third (control, n=27) or fourth Covid-19 vaccine dose in participants with chronic kidney disease (with and without organ transplant) (n=20), chronic kidney disease and organ transplant (n=16), immunosuppressive therapy (n=20) and no predisposing medical factors affecting the risk of Covid-19 (no risk group, n=17).

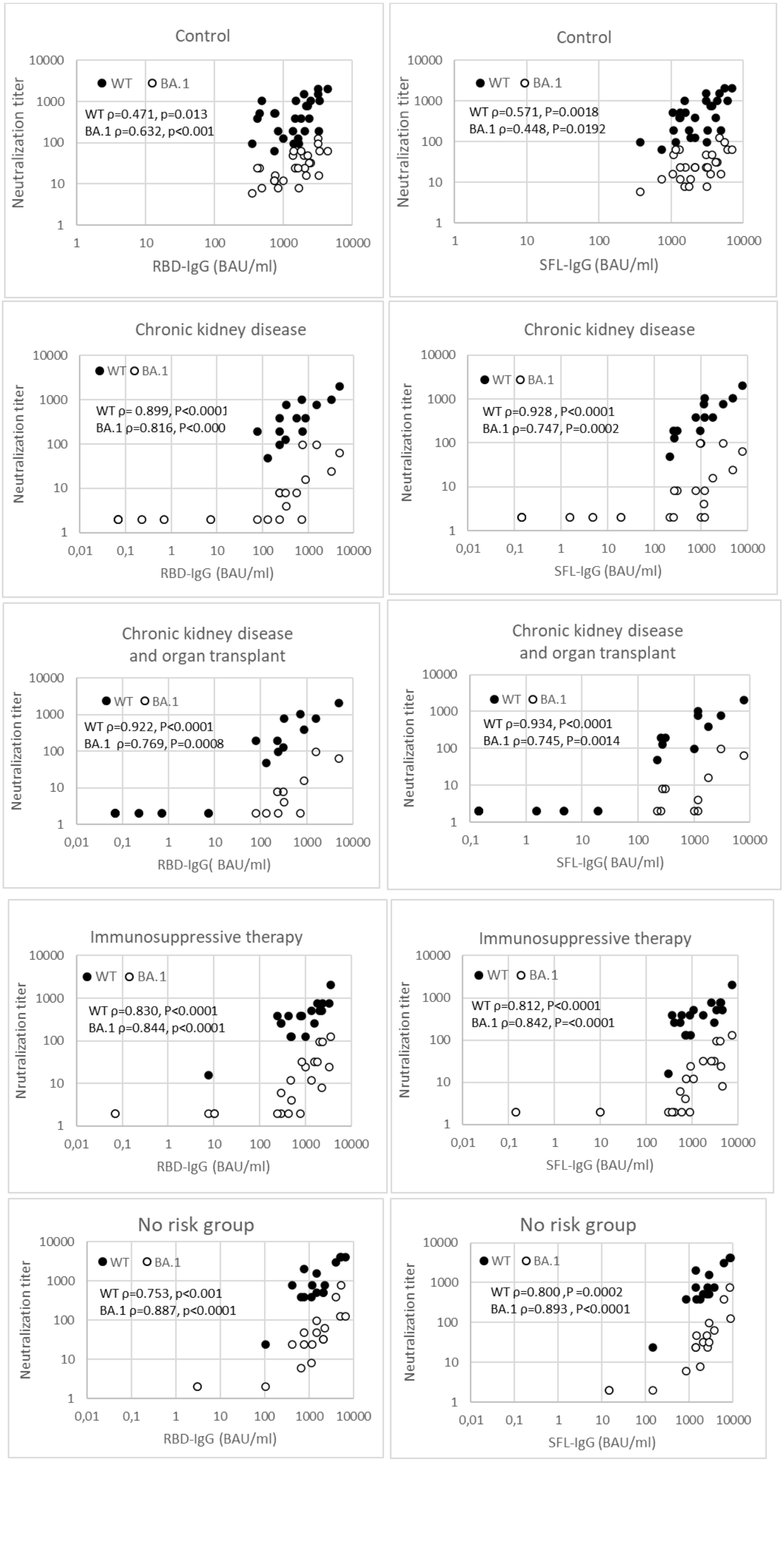

**Supplementary-Figure 2.** RBD-and SFL-IgG antibody concentrations (geometric mean with 95% confidence interval shown) one month after the fourth Covid-19 vaccine dose in participants with chronic kidney disease (CKD) and/or organ transplant. One participant can be in more than one group.

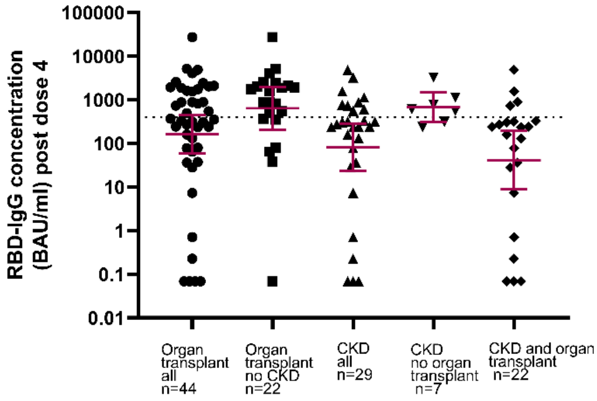


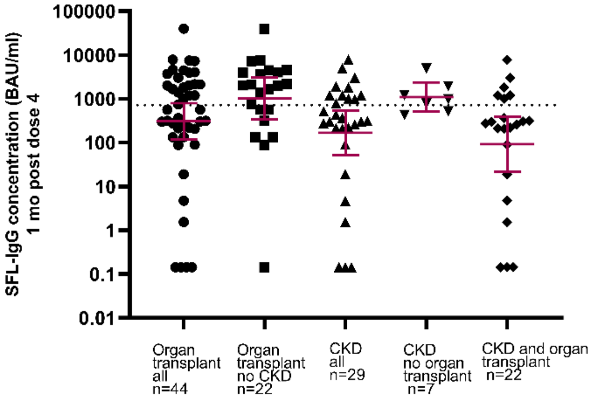
